# Priorities and barriers for research related to primary ciliary dyskinesia

**DOI:** 10.1101/2024.01.08.24300986

**Authors:** Myrofora Goutaki, Yin Ting Lam, Bruna Rubbo, James D Chalmers, Panayiotis Kouis, Gemma Marsh, Jean-François Papon, Johanna Raidt, Phil Robinson, Laura Behan, Jane S Lucas

**Affiliations:** Institute of Social and Preventive Medicine, University of Bern, Switzerland; Paediatric Respiratory Medicine, Children’s University Hospital of Bern, University of Bern, Switzerland; Primary Ciliary Dyskinesia Centre, University Hospital Southampton NHS Foundation trust, Southampton, UK; School of Clinical and Experimental Medicine, Faculty of Medicine, University of Southampton, Southampton, UK; Division of Molecular and Clinical Medicine, School of Medicine, University of Dundee, Dundee, UK; Respiratory Physiology Laboratory, Medical School, University of Cyprus, Nicosia, Cyprus; Children’s Therapy Department, Dorset County Hospital, Dorchester, UK; Depts of Paediatrics and Paediatric Respiratory Medicine, Imperial College and Royal Brompton Hospital, London, UK; Assistance Publique-Hôpitaux de Paris (AP-HP), Université Paris-Saclay, Hôpital Bicêtre, Service d’ORL, Le Kremlin-Bicêtre, France; Faculté de Médecine, Université Paris-Saclay, Le Kremlin-Bicêtre, France; Department of General Pediatrics, University Hospital Muenster, Muenster, Germany; Department of Respiratory and Sleep Medicine, Royal Children’s Hospital, Parkville, Australia; Department of Paediatrics, University of Melbourne, Parkville, Australia; Faculty of Medicine, University of Southampton, Southampton, UK

**Keywords:** Primary Ciliary Dyskinesia, orphan disease, research priorities

## Abstract

**Background:** Despite advances in primary ciliary dyskinesia (PCD) research, many questions remain; diagnosis is complex and no disease specific therapies exist. Using a mixed-methods approach, we aimed to identify priorities for clinical and epidemiological research and explore barriers to research.

**Methods:** To obtain rich, relevant, diverse data, we performed in-depth semi-structured interviews with PCD specialists selected using purposive sampling. We transcribed, coded, and analysed interview data using thematic analysis. Based on interview themes we identified, we developed an anonymous survey and circulated it widely through the BEAT-PCD network.

**Results:** We interviewed 28 participants from 15 countries across different disciplines and expertise levels. The main themes identified as priorities for PCD research were improving diagnosis, understanding prevalence, and disease course; phenotypic variability; disease monitoring; treatment strategies; clinical trial endpoints; and neglected research areas. In total, 136 participants (49% paediatric pulmonologists) from 36 countries completed the survey. Most commonly reported barriers for research were low awareness about PCD and difficulties securing funding—in more than one-third of cases, participants reported undertaking predominantly unfunded research. Research questions ranked highest included priorities related to further improving diagnosis, treating PCD, managing upper and lower airway problems, and studying clinical variability and disease prognosis.

**Conclusion:** We need to overcome barriers of limited funding and low awareness and promote collaborations between centres, disciplines, experts, and patients to address PCD priorities effectively. Our results contribute to the ongoing efforts of guiding the use of existing limited research resources and setting up a roadmap for future research activities.

**Take home message:** Our study defined PCD research priorities including improving diagnosis, treatments, managing upper and lower airway disease, and understanding prognosis. Key barriers identified include low disease awareness and limited funding opportunities.

## INTRODUCTION

Many factors hinder clinical and epidemiological research about rare diseases [1]. Low patient numbers even in reference centres, low awareness among clinicians and the public, and limited funding are among rare disease research barriers. In recent years, rare diseases became a health priority in Europe with initiatives such as the International Rare Diseases Consortium Initiative and the European Reference Networks (ERNs) [2, 3]. Supported by recent policies, rare lung disease research experienced unprecedented growth through collaborative efforts in Europe and abroad. ERN-LUNG was established in 2017 and focuses on several rare lung diseases [4, 5]. At the same time, the European Respiratory Society (ERS) supports several clinical research collaborations for developing large specific rare lung disease networks, including children’s interstitial lung disease, alpha-1 antitrypsin deficiency, and primary ciliary dyskinesia (PCD)[6–8]. In the field of PCD specifically, several research collaborations between clinicians and scientists have led to great advances understanding PCD better and improving patient care [9–12]. BEAT-PCD is a large international network set up initially in 2014 as a European Cooperation in Science and Technology (COST) Action (BM1407), which expanded in 2020 into an ERS-supported clinical research collaboration [8, 13]. The BEAT-PCD network provides an excellent opportunity to advance basic, clinical, and epidemiologic research and develop collaborative projects by building upon existing knowledge and utilising existing data resources [14–19]. Despite advances from recent years, PCD remains a neglected disease worldwide. PCD diagnosis improved but is still complex and varies significantly between and within countries. Since scant high-quality evidence supports development of PCD-specific guidelines, follow-up and management remain extrapolated from other diseases, such as cystic fibrosis or bronchiectasis [20–24]. Identifying research priorities, as well as potential challenges performing research on PCD supports the work of collaborative initiatives and research teams worldwide and guides the use of limited existing resources. Within the BEAT-PCD framework, our mixed-method study aimed to identify research gaps and priorities in clinical and epidemiological research in the field of PCD and explore barriers in research.

## METHODS

Our mixed-method study consisted of two parts: 1) a series of in-depth, semi-structured interviews with purposive selected healthcare professionals and researchers involved in PCD research and clinical care and 2) an anonymous electronic survey informed by interview findings, circulated widely through the BEAT-PCD network. The study received approval by the Faculty of Medicine Ethics Committee of the University of Southampton (ERGO 47010.A1).

### In-depth interviews

To obtain to rich, relevant, diverse data, we performed in-depth, semi-structured interviews with specialists involved in PCD research and clinical care selected using purposive sampling [25]. We invited and included participants from different countries, diverse research backgrounds, and various research experience levels (both senior and early career researchers). We aimed to interview participants from countries (participating in the BEAT-PCD network) with extensive, average, and limited research resources. Based on our aim, we mainly selected researchers with clinical management or clinical or epidemiological research experience; however, we also recruited basic scientists and diagnostic scientists to capture their opinions. All participants provided informed consent.

All interviews were conducted in English in-person or online between February 2019 and June 2021. Interviews followed a non-prescriptive guide developed in the beginning of the project and followed interviewee-raised issues opportunistically to ensure depth of information (Supplemental material). We derived interview guide questions from existing literature and feedback from collaborators. We tested them during the first 3–4 interviews, then adjusted the guide accordingly, adding prompts about issues raised by the first interviewees. The interviewer was the lead author (MG) who is full-time researcher in the field of PCD research and received in-depth interview and qualitative data analysis training for the project. With participant consent, interviews were audio-recorded and later transcribed verbatim by the lead (MG) or second author (YTL), in which case they were carefully validated by MG. Interview transcripts used an identification code to ensure and maintain interviewee anonymity; we removed identifying information from transcripts. We offered participants the opportunity to review the full transcript, upon request.

The planned sample size for the interviews was pragmatic; we based our estimations of a sufficient number for information power to reach breadth of information and collect rich, relevant, detailed data [26]. As with other rare diseases, the field of PCD research is limited and numbers of eligible participants restricted. Our objective involved collecting data to develop the survey by capturing different opinions about PCD research priorities, not achieving data saturation. MG coded and analysed data using an inductive thematic analysis approach [27, 28]. We grouped interview data in the subsequent coding steps until we identified common themes. We used NVivo software (1.5.2) to transcribe and analyse data; we followed the consolidated criteria for reporting qualitative research (COREQ) 32-item checklist for interviews and focus groups [29].

### Survey

Based on themes we identified from our interview data analysis, we developed a 21-question survey—taking an average 15 minutes to complete—including questions about a) general demographic, general participant PCD involvement, and specific PCD research; b) research funding for PCD projects and barriers for research; and c) research priority rankings grouped by main topics (diagnosis, presentation/prognosis, and follow-up, treatments, and other priorities) and overall. A multidisciplinary group of experts contributed to refining the survey questions by providing input on i) general question content, questions related to acquiring funding, and barriers for research and ii) wording and structuring of research priority questions. We developed the survey in English and programmed it in a Research Electronic Data Capture (REDCap) database hosted at the University of Bern (Supplementary material) [30]. We circulated study information about the survey via email to the BEAT-PCD network together with an invitation to participate. The BEAT-PCD mailing list includes more than 500 email addresses worldwide generally interested in PCD and our activities. We asked interested healthcare professionals and researchers contact the study team and consent to participate, then they received a link to complete the survey, which remained open between June 1–July 31, 2023. During this period, we sent two study reminders to the network. Survey participation was anonymous.

We presented survey results using descriptive statistics. For the overall priority ranking of research questions, we used a reciprocal ranking scoring system; each question received points based on its first (1 point), second (1/2 points), or third (1/3 points) overall research priority rank and 0 points if it was not ranked among the top three. Based on the final score, each question ranged from 0 to 1 point, with a higher score indicating a higher priority. We prioritized questions from highest to lowest mean score. We analysed the survey data using STATA version 15.1.

## RESULTS

### In-depth interviews

We interviewed 28 participants from 15 countries, 6 from outside Europe. Participants included 15 paediatric pulmonologists (several also cared for adult patients), 3 adult pulmonologists, 3 Ear-Nose-Throat (ENT) specialists, 1 specialist nurse, 1 physiotherapist, 1 epidemiologist, and 4 diagnostic scientists. All participants were involved in PCD care or research; not all were employed in specialist centres. Mean interview duration was 42 minutes and ranged from 18–77 minutes. Thirteen interviews took place face-to-face and 15 occurred online, using teleconferencing software.

During interviews, participants discussed their experiences with PCD-related research, barriers to successfully obtain funding, other factors that hinder or facilitate research on PCD in their institution and country, and the importance of collaborations and patient involvement in research. They also discussed their personal research interests in the field and expanded upon existing research gaps and priorities for future research. Although some participants mainly discussed high-level priorities, most expressed their preferences for specific questions addressed in the near future (Supplementary Table S1).

From interviews, the main themes we identified as important focal areas for clinical and epidemiological PCD research included 1) improving diagnosis, 2) prevalence and disease course, 3) phenotypic variability, 4) improving disease monitoring, 5) treatment strategies, 6) endpoints for clinical trials and research, 7) neglected areas, such as ENT and fertility problems and mental health issues, and 8) research in other ciliopathies and specific patient groups (Figure 1). We grouped priorities not directly linked to themes as other, more general priorities. We include representative quotes from interviews in Table 1.

**Figure 1:**
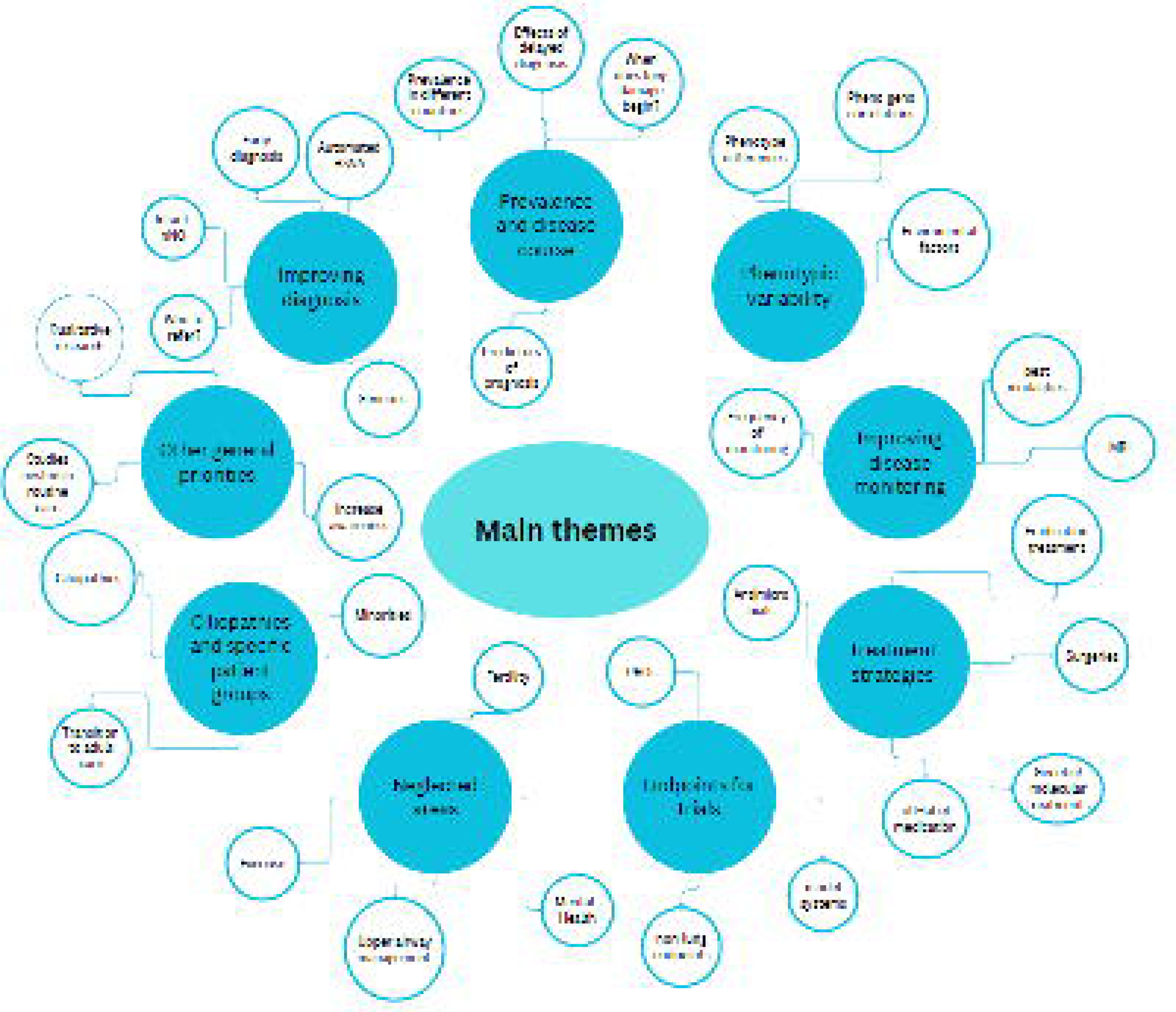
Main themes identified during in-depth interview by participants as important focus areas for clinical and epidemiological PCD research.

**Table 1:** Representative quotes related to PCD research from in-depth interviews with healthcare professionals and researchers.

| Topic | Quote |
| --- | --- |
| Challenges in PCD research | "I think the first challenge is just getting enough patients for [an] effective study. The second challenge is to fund, have enough funds for [a] coordinator, who can keep track of who is where and how to get in touch with them, get them in for new studies, so just the infrastructure. And then because of that you need to have to network with other centres. So [...] just finding other groups who share the same passion. I think everybody who does PCD research is probably under resourced and it's what their passion is, (laughs) that's why they do it, so you have to find other people who have the same passion. They are not in it for the money; that's for sure." |
| Lack of disease awareness | "[...] even [among] the health professionals there is no awareness and there is even ignorance many times, while there might be a clear suspicion for the diagnosis and it is clear that the next step towards the one or the other direction must be made it doesn't happen because simply, they do not have the knowledge." |
| Lack of awareness and interest from clinical community | "In general, physicians they have like one, maybe 2, maybe three PCD patients. So, it's difficult for them you know to be involved to think it's important." |
| Prioritising research topics | "Because just doing research for the sake of research is not as important as doing things that specifically people will find useful." |
| Research related to adult patients | "[We need more research] on adult PCD patients. We need to understand what this disease becomes; we need to understand the adult issues with complicated lung infections, fertility, things like that. It's been too paediatric based thus far, that's it." |
| Research about neglected areas | "I think fertility is a main issue for adult patients. I've seen so many young men and women and see that it was very difficult for them to find a good fertility expert. It takes time [...] So sometimes they miss the point, and they are not able to have children [...]." |
| Research about treatments | "I would like to [be able to] say [...] when I see a new patient where we could make the diagnosis [...] that yes, the evolution will be that [...] and then to say that there are a few treatments, validated by studies." |
| Integrated research approach | "I think historically, clinical researchers focus on a single area, you know, so they come at these diseases from a microbiological perspective, or they come at these diseases from an immunology perspective. And I think all of those are [...] that approach is quite flawed because these diseases are highly complex. So if you're only interested in diagnostics, or if you're only interested in microbiology, you are always I think, hitting up against the barriers of the questions you can answer." |
| Research collaborations | "I think the only solution would be collaboration because [...] nobody can get money for everything. So I think there must be a focus. Some research groups have to have different focuses, so that it might be more possible to get funding for this. Because when everybody wants to have funding for the same things, it could be difficult. I think you need to collaborate and as we did in the BEAT-PCD also, you have to get in groups and then maybe funding is more possible than if you were just playing alone." |
| Patient involvement in research | “Rather than just recruiting patients into a study, we should much more have equal partnerships with people who have the disease, already from the start of a study, for designing a study, for designing questionnaires, for designing approaches, how to inform people with PCD about the study and how to participate, how to interpret results.” |

We edited quotes for brevity, clarity, and correctness; we present quotes as mostly verbatim, while also ensuring anonymity.

### Survey-barriers for PCD research

We excluded 4 (3 incomplete; 1 duplicate) of 140 filled-in questionnaires. Participants represented 36 countries and 63% were female (Table 2). Most were paediatric pulmonologists (49%), followed by ENT specialists (10%), diagnostic scientists (9%), adult pulmonologists (7%), paediatricians (7%), other healthcare professionals (6%), namely specialist nurses and physiotherapists, epidemiologists, or data scientists (5%), and other (7%) (Table 2). Of survey respondents, 81% reported involvement with care or diagnosis of patients with PCD and 70% involvement in research currently; overall 82% reported personal involvement in PCD research. Almost half (47%) reported experience with PCD >10 years; 31% 5–10 years; and 22% <5 years (Table 2).

**Table 2:** Characteristics of participants in the online survey about priorities and barriers for PCD research (N=136).

|  | **Total N (%)** |
| --- | --- |
| **Female sex** | 86 (63) |
| **Country of residence** |  |
| United Kingdom | 18 (13) |
| Switzerland | 14 (11) |
| Turkey | 13 (10) |
| Spain | 11 (8) |
| Germany | 8 (6) |
| France | 7 (5) |
| Israel | 6 (4) |
| Italy | 6 (4) |
| United States | 6 (4) |
| Other European countries | 28 (21) |
| Other non-European countries | 19 (14) |
| **Involvement with PCD** |  |
| Research and diagnosis or care | 76 (56) |
| Diagnosis or care | 34 (25) |
| Research | 19 (14) |
| Other | 7 (5) |
| **Occupation** |  |
| Paediatric pulmonologist | 69 (51) |
| Adult pulmonologist | 9 (7) |
| ENT specialist | 14 (10) |
| Diagnostic scientist | 12 (9) |
| Paediatrician | 9 (7) |
| Non-physician healthcare professional | 8 (6) |
| Epidemiologist or data scientist | 7 (5) |
| Other | 8 (6) |
| **Work setting** |  |
| Academic hospital | 102 (75) |
| Academic research institution | 21 (15) |
| Non-academic hospital | 9 (7) |
| Other | 4 (3) |
| **Years of involvement in PCD research or care** |  |
| >10 years | 64 (47) |
| 5–10 years | 42 (31) |
| <5 years | 30 (22) |
| **Participated in PCD research during the past 15 years** | 112 (82) |

PCD: primary ciliary dyskinesia; ENT: ear, nose, and throat. Characteristics are presented as N and column %; due to rounding, numbers do not always add up to 100%.

Survey respondents reported PCD research as most usually funded by competitive research grants, institutions or governments, and smaller foundations (Table 3) or as unfunded (35%). Nearly half (49%) previously applied for PCD research funding and over half (51%) considered funding for PCD more difficult to acquire than for other diseases. Respondents reported the most important barriers for acquiring funding included low awareness about PCD, high competition for funding, lack of commercial application, and rarity of the disease. Other factors they reported as hindering PCD research involved lack of dedicated research time (68%), small numbers of patients in each centre (63%), inactive or non-existent patient support groups (63%), disease heterogeneity (58%), few colleagues with expertise in PCD locally (57%), and lack of needed resources such as specialised equipment or databases (46%) (Table 3). Participants strongly agreed about the need for national and international multidisciplinary research collaborations (82%), the importance of registries and cohort studies as research tools (89%), the importance of patient support groups (76%), and actively involving patients in different stages of research (78%), as well as the need for standardised care and information collection to improve future data quality (85%).

**Table 3:** Barriers and factors facilitating clinical and epidemiological research related to PCD according to the online survey participants (N=136)

|  | Total N (%) |
| --- | --- |
| **Main source of funding for PCD research<sup>¶</sup>** |  |
| Institutional/ governmental funding | 56 (41) |
| Competitive grants | 57 (42) |
| Funding from smaller foundations | 41 (30) |
| Funding from collaborative research | 24 (18) |
| Unfunded | 48 (35) |
| **Compared to other diseases in the field, obtaining funding for PCD is** |  |
| More difficult | 69 (51) |
| Easier | 1 (1) |
| No difference | 29 (21) |
| I do not know | 37 (27) |
| **Barriers in acquiring funding for PCD clinical and epidemiological research<sup>#</sup>** |  |
| Low awareness about PCD | 113 (83) |
| High competition for funding | 102 (75) |
| Lack of commercial application | 89 (65) |
| Rarity of disease | 80 (59) |
| Low mortality rate/ not considered severe | 58 (43) |
| Lack of supporting evidence/ existing research framework | 57 (42) |
| Lack of local support in preparing a funding application | 54 (40) |
| Lack of expertise of research team/ limited publication record | 44 (32) |
| Higher interest in basic research projects | 41 (30) |
| **Other factors hindering PCD research<sup>#</sup>** |  |
| Lack of dedicated research time | 93 (68) |
| Small numbers of patients | 86 (63) |
| No/inactive patient support group | 86 (63) |
| Disease heterogeneity | 79 (58) |
| Few colleagues locally with expertise in PCD | 78 (57) |
| Lack of needed resources | 64 (47) |
| Lack of good local or extended collaborative network | 38 (28) |
| Lack of interest about PCD from most colleagues | 38 (28) |
| Lack of motivation to participate from patients | 31 (23) |
| **Factors facilitating PCD research<sup>#</sup>** |  |
| National and international registries and cohort studies | 121 (89) |
| National and international multidisciplinary collaborations | 112 (82) |
| Standardisation of care and collected information to improve data quality | 116 (85) |
| Active involvement of patients in research | 106 (78) |
| Patient support groups | 103 (76) |

PCD: primary ciliary dyskinesia. y: years. Characteristics are presented as N and column %, ¶: multiple answers possible, #: 5-point Likert scale responses, here presented are agree or strongly agree

### Survey-priorities for PCD research

Nearly all participants (135/136) responded to the research priority ranking questions. Among the three questions related to PCD diagnosis, nearly half of participants (49%) chose “How to improve the accuracy, speed, and cost-effectiveness of diagnostic testing in different age groups and health care settings?” (Figure 2) as the most important question. In the questions related to PCD presentation, prognosis, and follow-up, most participants selected “What is the clinical variability and natural course of upper and lower respiratory disease in PCD and which factors affect disease prognosis?” (41%) and “Which health-related behaviours or everyday interventions can have a positive role in improving symptoms or quality of life in people with PCD?” (27%) as the most important questions. The most important questions for participants regarding PCD treatments asked, “Are there any genetic or molecular treatments in the pipeline that could help restoring ciliary function?” (33%) and “Which of the already available and currently used medication and other management approaches for upper and lower airways are suitable for PCD patients?” (29%), respectively. Among other research priorities, increasing awareness and engagement of clinicians and patients in PCD research (54%) was reported as most important.

**Figure 2:**
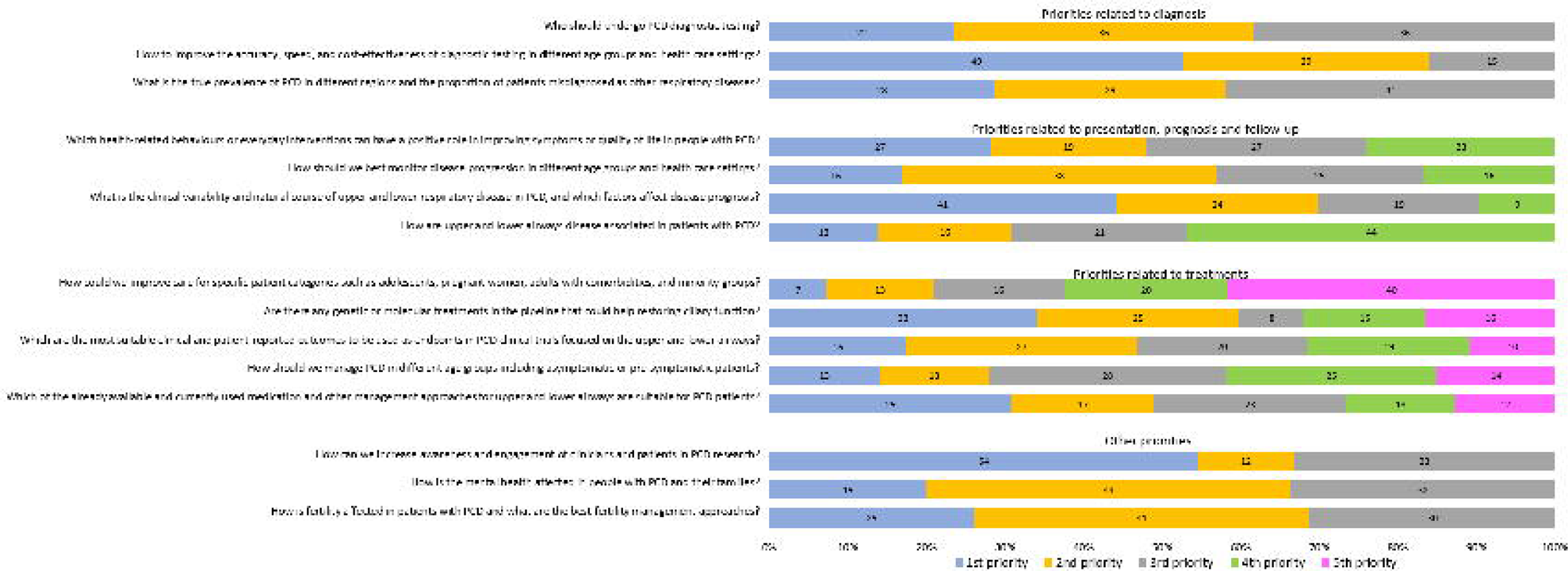
Ranked research priority questions by survey participants grouped by main topic.

Ranked survey participant opinions on overall top priorities—across all topics related to PCD clinical and epidemiological research—varied with scores ranging from 0.02 to 0.31 (Table 4).

**Table 4:** Top priorities across all topics related to PCD clinical and epidemiological research as ranked by survey participants.

| Rank | Top priorities across all topics related to PCD clinical and epidemiological research | Mean score |
| --- | --- | --- |
| 1 | How to improve the accuracy, speed, and cost-effectiveness of diagnostic testing in different age groups and health care settings? | 0.312 |
| 2 | Are there any genetic or molecular treatments in the pipeline that could help restoring ciliary function? | 0.273 |
| 3 | What is the clinical variability and natural course of upper and lower respiratory disease in PCD, and which factors affect disease prognosis? | 0.199 |
| 4 | Which of the already available and currently used medication and other management approaches for upper and lower airways are suitable for PCD patients? | 0.169 |
| 5 | Who should undergo diagnostic testing? | 0.168 |
| 6 | What is the true prevalence of PCD in different regions and the proportion of patients misdiagnosed as other respiratory diseases? | 0.121 |
| 7 | Which are the most suitable clinical and patient-reported outcomes to be used as endpoints in PCD clinical trials focused on the upper and lower airways? | 0.115 |
| 8 | Which health-related behaviours or everyday interventions can have a positive role in improving symptoms or quality of life in people with PCD? | 0.114 |
| 9 | How should we best monitor disease progression in different age groups and health care settings? | 0.101 |
| 10 | How can we increase awareness and engagement of clinicians and patients in PCD research? | 0.077 |
| 11 | How should we manage PCD in different age groups including asymptomatic or pre-symptomatic patients? | 0.070 |
| 12 | How is the mental health affected in people with PCD and their families? | 0.028 |
| 13 | How is fertility affected in patients with PCD and what are the best fertility management approaches? | 0.025 |
| 14 | How could we improve care for specific patient categories such as adolescents, pregnant women, adults with comorbidities, and minority groups? | 0.023 |
| 15 | How are upper and lower airways disease associated in patients with PCD? | 0.021 |

Questions ranked from most to least important based on the mean of a reciprocal ranking score (0–1); each question ranked either first (1 point); second (1/2 points); third (1/3 points), or not ranked (0 points) among the top 3 priorities.
PCD: primary ciliary dyskinesia

The three questions asked, “How to improve the accuracy, speed, and cost-effectiveness of diagnostic testing in different age groups and health care settings?” (ranked first); “Are there any genetic or molecular treatments in the pipeline that could help restoring ciliary function?” (second); and “What is the clinical variability and natural course of upper and lower respiratory disease in PCD, and which factors affect disease prognosis?” (third; Text Box 1). All three questions related to PCD diagnosis ranked in the top six of the overall priorities list. Questions related to relatively neglected areas from a research perspective, such as upper airways, mental health, fertility, and care for specific patient groups and minorities, ranked lowest among participants (Table 4). When comparing rankings between paediatric pulmonologists (largest groups) and other specialties, top priorities remained the same; however, results showed differences among lower ranked priorities (Supplementary Table S2).

#### Text Box 1: Top 3 research questions for PCD clinical and epidemiological research according to healthcare professionals and researchers

- How to improve the accuracy, speed, and cost-effectiveness of diagnostic testing in different age groups and health care settings?
- Are there any genetic or molecular treatments in the pipeline that could help restoring ciliary function?
- What is the clinical variability and natural course of upper and lower respiratory disease

## DISCUSSION

Using a mixed-method approach, our study identified main priorities and explored opinions about barriers for clinical and epidemiological research related to PCD as perceived by PCD professionals and researchers. Research in rare diseases, such as PCD, often faces different and additional challenges compared with more common conditions [31, 32]. By identifying specific barriers and factors for facilitating PCD-related research, it supports researchers and strengthens efforts to address these difficulties. Furthermore, when developing future research agendas, our results provide a roadmap for BEAT-PCD and the PCD community overall.

Our study’s main strengths include the mixed-methods design and far reach of the BEAT-PCD network [33]. Employing qualitative methods allowed us to gain rich information about healthcare professional and researcher perspectives, from participants from various background, areas and time of expertise, which we then used as a basis to develop a widely circulated survey [34–36]. Our approach ensured we noticed important aspects and allowed discussions of perspectives from purposefully selected participants in more detail than from questionnaires alone. Since most interviewees were prior acquaintances of MG through the BEAT-PCD network, it created an environment of trust and comfort for open discussions. Yet, the familiarity possibly influenced participant answers during the interviews [37]. Through the BEAT-PCD mailing list we widely distributed information about the study and invitations for survey participation. However, since the mailing list includes people generally interested in PCD and our activities—some recipients were likely ineligible for participation, such as patient representatives or healthcare professionals with very limited experience in the field—it was not possible to calculate a survey response rate.

The main limitation of the study involves survey respondents closely representing the distributions of country, discipline, and experience level with PCD in the BEAT-PCD network. We accomplished representation of experts from many countries with organised PCD care and research activities and high participation numbers among paediatric pulmonologists. Our participants included fewer participants from other specialties, such as adult pulmonologists, ENT specialists, and other healthcare professionals, which highlights a need to increase multidisciplinary collaborations and awareness in other fields. Since paediatric pulmonologists represented only half of survey participants, it is noteworthy our study results include perspectives from other disciplines. Our study deliberately focused on healthcare professionals and researchers; we did not include people with PCD or parents of affected children—a separate, dedicated study focusing on patient and family perspectives regarding PCD research is ongoing [38]. Another limitation includes only one person coding all interviews. Although we followed an inductive approach, thematic analysis often relies on researcher judgement, possibly introducing biases from their own interpretations [37]. MG is a female, clinical epidemiologist with extensive experience in the field of epidemiological and clinical PCD research; she currently co-chairs the BEAT-PCD network. She coded and analysed the interview data under this lens. Part of the interviews were completed pre-pandemic, while others during the COVID19 pandemic; however analysis did not show any evident difference in themes.

Most commonly reported barriers for PCD research were low awareness about the disease, which has already been previously reported, and difficulties in securing funding[39, 40]. Half of participants considered obtaining funding for PCD more difficult when compared with other diseases in the field; in more than one-third of cases, research was mostly performed without funding. Ongoing efforts from past years—promoting multidisciplinary collaborations, exchanging expertise, and sharing resources, such as setting up registries and multicentre cohort studies—are all steps in the right direction to address these barriers, yet space remains for further improvement.

Highlighted by relatively low mean ranking scores even for top-ranked priorities, we found variability in research question priority ranking. The finding emphasizes several enduring important research gaps, instead of a clear consensus on just a handful of major priorities for PCD research. Participants from different disciplines possess different interests, which possibly reflects ranking. Top-ranked priorities were related to further improving diagnosis; treating PCD and managing upper and lower airway problems; and studying clinical variability and disease prognosis—all questions considered unlinked to specific disciplines. Our findings become more meaningful in the light of ongoing efforts to develop joint updated diagnostic guidelines for PCD; new potential molecular treatments in the pipeline; and the development of the PCD-specific clinical trials network [41, 42]. Notably, other topics strongly impacting the lives of people with PCD and their families, such as fertility or mental health, ranked lower by experts, although they appeared in the priority list. In a study of bronchiectasis research priorities—in addition to topics included in the expert consensus—42% of patient participants outlined additional topics, including research related to mental health [43]. For α1-antitrypsin deficiency, patients and caregivers included development of other aspects of integral care, such as caregiver support and psychological care, as their most important research areas, while respiratory specialists did not [44]. To ensure a common direction for the PCD research community with patient support groups and affected individuals, comparing priorities of people affected with PCD is an important next step [45].

Our study is the first assessing priorities and barriers for PCD research; it combines rich and detailed perspectives from in-depth interviews and representative high-level information from the PCD research community. We need to overcome barriers of limited funding and low disease awareness and promote collaborations between centres, disciplines, experts, and patients to address priorities effectively. Our results contribute to ongoing efforts to guide the use of existing, limited research resources and setting up a roadmap for future research activities to improve and streamline research in the field.

## Data Availability

All data produced in the present study are available upon reasonable request to the authors.

## Author Contributions

M Goutaki and J. S Lucas developed the concept and designed the study. M Goutaki, J. S Lucas, and L Behan developed the interview guide. M Goutaki conducted in-depth interviews and M Goutaki and YT Lam transcribed them. M Goutaki performed thematic analysis, developed the survey, cleaned the dataset, performed the statistical analyses, and drafted the manuscript. All authors commented and revised the survey and manuscript. M Goutaki takes final responsibility for content.

## Funding

We acknowledge the support of the European Respiratory Society (ERS) Short-Term Research fellowship Nr 201804-00367. The study was developed as a collaboration between the Universities of Southampton and Bern within the framework of the BEAT-PCD COST Action BM1407 and completed in the BEAT-PCD clinical research collaboration framework, supported by ERS. M Goutaki is supported by a Swiss National Science Foundation Ambizione fellowship (PZ00P3_185923) and the Johanna Dürmüller-Bol foundation. Several authors participate in ERN-LUNG (PCD core).

### Acknowledgments

We thank all healthcare professionals and researchers who participated in the in-depth interviews or completed the survey. We thank Corine M. Driessens, Mary E. Barker, and Polly Hardy-Johnson (University of Southampton) for their advice and support developing the interview guide and thematic analysis and Kristin Marie Bivens (University of Bern) for her editorial assistance.

## Conflicts of Interest

We declare no conflicts of interest.

## Notes

### Competing Interest Statement

The authors have declared no competing interest.

### Author Declarations

The Faculty of Medicine Ethics Committee of the University of Southampton gave ethical approval for this work(ERGO 47010.A1).

