## Supplementary Table S1 for "Priorities and barriers for research related to primary ciliary dyskinesia"

**Supplementary table S1:** Specific research questions on different topics related to PCD research, identified during the in-depth interviews with selected experts.

**Diagnosis**

1. How can we diagnose PCD more accurately in different settings/countries?
2. How can we improve efficiency and affordability of PCD diagnosis in settings with limited resources (financial or expertise-wise)?
3. Does increasing awareness about PCD in general practitioners improve referral of people suspected for PCD and their further diagnosis?
4. When should non-respiratory specialists (e.g. ENT or fertility specialists) refer a patient for PCD diagnostic work-up?
5. How can nNO be used as a screening tool to improve referral of people suspected to have PCD for diagnosis?
6. Which groups of people with PCD have normal nNO and why?
7. What is the role of nNO measurement in diagnosis of PCD in infancy?
8. How can we improve diagnosis of PCD in the neonatal period?
9. How can the use of automated procedures/artificial intelligence improve the diagnosis of PCD?
10. Can we further improve the value of genetic testing for PCD diagnosis?
11. How can we standardise all diagnostic tests for PCD worldwide?
12. Are there any specific characteristics of common symptoms like cough or rhinitis that are more relevant/indicative of PCD?

**Prevalence/disease course- prognosis**

13. What is the prevalence of PCD in different countries/regions?
14. What is the impact of delayed diagnosis/misdiagnosis on disease course of PCD?
15. What proportion of adults with idiopathic bronchiectasis or other common adult respiratory diagnosis actually have undiagnosed PCD?
16. How early in life does lung damage start in PCD?
17. How do symptoms of people with PCD change with seasons?
18. Which baseline characteristics could be used to predict prognosis of PCD?
19. What is the role of the microbiome in PCD disease progression?
20. Which factors that we can influence contribute to disease prognosis for PCD?
21. What is the role of non-tuberculous mycobacteria in chronic infections in people with PCD?

**Clinical variability/ genotype-phenotype correlations**

22. What are the specific characteristics that differentiate clinical phenotypes in PCD?
23. Which genes/ultrastructural defects are associated with better prognosis in PCD?
24. Which are the determinants of clinical variability and disease severity in PCD?
25. Are PCD phenotypes only based on genetics or do environmental, socio-economic factors and health-related behaviours play a role in phenotypic variability?
26. Which factors could explain the disease variability between siblings with the same PCD- causing genetic mutation?

**Disease monitoring**

27. What is the role of regular and functional MRI for monitoring PCD disease course?
28. Which are the best modalities to assess early lung disease in people with PCD?
29. What is the correlation between imaging and lung function in PCD?
30. What is the best frequency of monitoring of people with PCD and with which tests?
31. How can we standardise lung function monitoring in people with PCD?
32. Which are the best modalities to monitor lung function in people with PCD of different ages?

**Treatment strategies**

33. When and how should we treat microbial colonisation of the lungs and the upper airways in people with PCD?
34. What is the best eradication treatment approaches in PCD?
35. What is the role of antimicrobial treatments in management of people with PCD?
36. What is the role of anti-inflammatory medication in the management of people with PCD?
37. What are the best measures to avoid cross-infections for people with PCD?
38. What is the best way to treat complicated lung infections in adults with PCD?
39. What is the role of anti-neutrophil treatments in management of people with PCD?
40. Is there a role for targeted antibodies against pathogens for the management of people with PCD?
41. What is the role of recombinant DNase in care of people with PCD?
42. Which are the indications for surgical management (lungs and upper airways) for people with PCD?
43. Which are the most effective physiotherapy techniques to reduce symptoms and improve quality of life in people with PCD?
44. Which treatments used in management of other respiratory diseases work equally well or better in the management of PCD?
45. What are the complications of off-label medication used in management of PCD?
46. What is the role of personalised-management approaches in PCD?
47. How should we manage asymptomatic/pre-symptomatic PCD?
48. How should we care for adults with PCD and other underlying health conditions?
49. How can we delay the onset of bronchiectasis in people with PCD diagnosed in childhood?

**Clinical trials/ trial endpoints**

50. Which are the most suitable non-lung related endpoints relevant for clinical trials in PCD?
51. Which are the most suitable patient-reported outcomes to be used as endpoints in clinical trials in PCD?
52. Is there a place for pragmatic trials in generating evidence for PCD management?
53. How can we improve evidence for the efficacy of off-label medication used in people with PCD?
54. How can we best standardise clinical measures for use in research and randomised trials?
55. How can management of PCD be standardised worldwide?
56. Which genetic/molecular treatments could help restoring ciliary function?
57. Which medication could improve mucociliary clearance?
58. How can we improve existing model systems available for PCD for use in clinical trials?

**Fertility**

59. When should people with PCD be referred to fertility specialists?
60. How is the female fertility affected in PCD?
61. How are women with PCD affected in pregnancy and how can we support their management?
62. Which assisted fertility approaches are best for people with PCD?
63. What type of family counselling do people with PCD need?

**Upper airways**

64. How is upper respiratory disease associated with lower respiratory disease and especially pulmonary exacerbations?
65. What are the different elements of upper airway exacerbations compared to pulmonary exacerbations?
66. How do upper airway symptoms change throughout the disease course and how should management be adapted?
67. How do upper airway symptoms affect quality of life for people with PCD?
68. How does the pathophysiology of upper airway disease in PCD differ from other chronic ENT conditions?
69. What is the role of nasal rinsing in the management of chronic rhinosinusitis in people with PCD?
70. How should we best manage hearing impairment in people with PCD?
71. Do nasal steroids improve symptoms in people with PCD and rhinosinusitis?
72. In which cases should we treat recurrent otitis with ventilation tubes in people with PCD?

**Health-related behaviours/mental health**

73. What types of exercise are most beneficial for people with PCD?
74. Which health-related behaviours/everyday interventions can have a positive role in improving symptoms/quality of life in people with PCD?
75. How does PCD affect different aspects of daily lives of people with PCD?
76. How can we improve the mental health of people with PCD?
77. What is the treatment burden for people with PCD and their families?

**Ciliopathies**

78. Which disease characteristics are shared between PCD and other ciliopathies and how these affect their management?
79. Is there a respiratory involvement in other ciliopathies?

**Research on special groups/periods of life**

80. What aspects of PCD and its management are particularly relevant for minority groups?
81. How can we improve transition from paediatric to adult care?

**General priorities**

82. How can we improve/encourage research nested in routine clinical care?
83. How can we adopt successful integrated research approaches for PCD?
84. How can we improve therapeutic education for people with PCD and their families?
85. How can we improve adherence to treatment in people with PCD of different ages?
86. How do we raise awareness and knowledge of PCD in non-specialists pulmonologists and physicians of other specialties?
87. How do we raise the public’s awareness and knowledge of PCD?
88. How can we increase patient engagement in research?
89. How can research support nursing practices?
90. How can we best incorporate qualitative elements in PCD research?

PCD: primary ciliary dyskinesia, ENT: ear-nose-throat, nNO: nasal Nitric Oxide

**Supplementary table S2:** Top priorities across all topics related to PCD research as ranked by paediatric pulmonologists and other participants

| **Rank** | **Top priorities across all topics related to PCD clinical and epidemiological research** | **Mean score** | | |
| --- | --- | --- | --- | --- |
|  |  | **Total**  **(N=135)** | **Paed pulm**  **(N=67)** | **All other**  **(N=68)** |
| 1 | How to improve the accuracy, speed, and cost-effectiveness of diagnostic testing in different age groups and health care settings? | 0.312 | 0.348 | 0.277 |
| 2 | Are there any genetic or molecular treatments in the pipeline that could help restoring ciliary function? | 0.273 | 0.276 | 0.270 |
| 3 | What is the clinical variability and natural course of upper and lower respiratory disease in PCD, and which factors affect disease prognosis? | 0.199 | 0.201 | 0.196 |
| 4 | Which of the already available and currently used medication and other management approaches for upper and lower airways are suitable for PCD patients? | 0.169 | 0.164 | 0.174 |
| 5 | Who should undergo diagnostic testing? | 0.168 | 0.197 | 0.140 |
| 6 | What is the true prevalence of PCD in different regions and the proportion of patients misdiagnosed as other respiratory diseases? | 0.121 | 0.117 | 0.125 |
| 7 | Which are the most suitable clinical and patient-reported outcomes to be used as endpoints in PCD clinical trials focused on the upper and lower airways? | 0.115 | 0.110 | 0.118 |
| 8 | Which health-related behaviours or everyday interventions can have a positive role in improving symptoms or quality of life in people with PCD? | 0.114 | 0.060 | 0.152 |
| 9 | How should we best monitor disease progression in different age groups and health care settings? | 0.101 | 0.112 | 0.091 |
| 10 | How can we increase awareness and engagement of clinicians and patients in PCD research? | 0.077 | 0.102 | 0.081 |
| 11 | How should we manage PCD in different age groups including asymptomatic or pre-symptomatic patients? | 0.070 | 0.082 | 0.066 |
| 12 | How is the mental health affected in people with PCD and their families? | 0.028 | 0.035 | 0.020 |
| 13 | How is fertility affected in patients with PCD and what are the best fertility management approaches? | 0.025 | 0.010 | 0.039 |
| 14 | How could we improve care for specific patient categories such as adolescents, pregnant women, adults with comorbidities, and minority groups? | 0.023 | 0.005 | 0.042 |
| 15 | How are upper and lower airways disease associated in patients with PCD? | 0.021 | 0 | 0.042 |

Questions ranked from most to least important based on the mean of a reciprocal ranking score (0–1); each question ranked either first (1 point); second (1/2 points); third (1/3 points), or not ranked (0 points) among the top 3 priorities.

PCD: primary ciliary dyskinesia, Paed pulm: paediatric pulmonologists.
