## Supplemental material for "Priorities and barriers for research related to primary ciliary dyskinesia"

**Project title**: **Identifying knowledge gaps for primary ciliary dyskinesia research**

**Interview Guide**

The interviews will be conducted in a semi-structured way. The questions below are meant to serve as a guide; the researcher will adapt each interview according to the relevance of the different section to the interviewee. For example, for researchers working in countries with limited research resources, the interviewer might spend more time discussing difficulties in raising research funding and to which direction the available limited resources should focus. On the other hand, for researchers working in countries with more extensive resources and who have extensive experience in research, the interviews will focus on expanding research possibilities, utilizing new available resources and building up on current advances. This interview guide is not prescriptive, and issues raised by the interviewee will be followed opportunistically. The main objective of these interviews is to explore opinions of researchers on current and future PCD-related research in order to develop a survey that will be distributed to all BEAT-PCD members and which will be used to achieve prioritisation of the issues identified through the interviews and will measure how researchers agree with these opinions.

**Introduction**

Thank you very much for agreeing to participate in this study.

As I mention in the Participant Information Sheet, this study is a project within BEAT-PCD (COST Action BM1407), developed with the support of the European Respiratory Society. The aim of this interview is to gain a deeper understanding of the different knowledge gaps for clinical and epidemiological research and the importance of different research themes. This information will be used for the development of a survey on research priorities in the field which will be send to all BEAT-PCD participants. It is important that we design the survey in a way that is relevant to different settings, as we are fully aware that research resources might differ importantly between the countries that participate to the BEAT-PCD network.

The interview will take roughly an hour so I would like to confirm with you that we do have sufficient time for this. Obviously if anything urgent comes up during the interview please let me know and we can arrange a convenient time to resume the interview.

Do you have any questions for me before we start?

**Participants’ role**

These first few questions are important so we can understand your experiences and interests in PCD-related research.

1. Please describe your involvement with the field of PCD
2. How long have you been a member of the BEAT-PCD network?
3. In which BEAT-PCD activities have you been involved?
4. What % of your time you would say you are involved in research?
5. How would you describe your involvement to PCD-related research?
6. Please describe your personal research interests in PCD research.
7. Which are the specific areas of PCD research you are currently involved?
8. Please describe your role in the PCD- related research projects you are currently involved.

**Research resources**

I would like to ask you a few questions related to the research resources available in your centre/country.

1. Please describe how PCD research is funded in your centre/country.
2. Please expand on the difficulties to raise funding for PCD research in your centre/country.
3. Were there occasions when your efforts to get funding for PCD research were unsuccessful and if yes, could you elaborate on the main issues that were put forward by the funding bodies/reviewers?
4. Are there any additional challenges in performing PCD-related research in your centre/country?

**Opinions**

This set of questions focuses primarily in epidemiological and clinical PCD-related research. We would like to discuss your opinions about PCD-related research priorities and about how you see the future of research in this area.

1. What are your thoughts about the achievements of PCD research during the last few years?
2. Which do you think are the most important knowledge gaps currently in the field?
3. Which research questions do you think the PCD research community should try to tackle in the next few years?
4. Which PCD-related questions are you more interested in seeing answered in the near future based on your personal research interests?
5. What do you think about the involvement of patients/ patient organisations in PCD research?
6. What is your opinion about the role of collaborative research initiatives for PCD?
7. What are your thoughts regarding the relationship of research needs and available resources in the field of PCD?
8. What are your thoughts regarding data sharing and utilisation of available research resources from other research groups?
9. What do think about the role of national and multinational registries and patient cohorts in PCD-related research?
10. Are you aware of any available datasets and research resources, which could be utilized to research the issues you have highlighted as priority? Please expand on this.
11. How do you think we could improve the quality of available data for future research?
12. How do you think PCD-related research could utilize new emerging methodology and techniques to develop further in the following years?
