## Supplementary material for "Priorities and barriers for research related to primary ciliary dyskinesia"

You have been invited because of your experience on PCD research or clinical care of PCD patients.

Your participation will help us prioritize research needs for clinical and epidemiological research in the field and suggest future studies addressing these issues.

Please complete the survey below.

The survey is  anonymous, thus your data and participation are confidential.

Thank you!

---

Are you (select all that apply):

- ☐ Involved in diagnosis or care of patients with primary ciliary dyskinesia?
- ☐ Involved in primary ciliary dyskinesia research?
- ☐ Other

---

If other, please explain

__________________________________

---

Are you (select what applies best):

- ◯ A paediatric pulmonologist
- ◯ An adult pulmonologist
- ◯ An ENT specialist
- ◯ A paediatrician
- ◯ A general practitioner
- ◯ A physician of a different specialty than those listed above
- ◯ A diagnostic scientist
- ◯ An epidemiologist, data scientist, or bioinformatician
- ◯ A non-physician healthcare professional (e.g. nurse, physiotherapist, audiologist, physiologist, psychologist)
- ◯ Other

---

Which specialty?

__________________________________

---

Which is your occupation?

__________________________________

---

If other, please explain

__________________________________

---

In which health care or research setting are you employed (select what applies best)?

- ◯ Academic hospital
- ◯ Non-academic hospital (public or private)
- ◯ Private practice
- ◯ Academic research institution
- ◯ Private or industrial research unit
- ◯ Other

---

If other, please explain

__________________________________

In which country are you working?

◯ Afghanistan
◯ Albania
◯ Algeria
◯ Andorra
◯ Angola
◯ Antigua and Barbuda
◯ Argentina
◯ Armenia
◯ Australia
◯ Austria
◯ Azerbaijan
◯ Bahamas
◯ Bahrain
◯ Bangladesh
◯ Barbados
◯ Belarus
◯ Belgium
◯ Belize
◯ Benin
◯ Bhutan
◯ Bolivia
◯ Bosnia and Herzegovina
◯ Botswana
◯ Brazil
◯ Brunei
◯ Bulgaria
◯ Burkina Faso
◯ Burundi
◯ Cambodia
◯ Cameroon
◯ Canada
◯ Cape Verde
◯ Central African Republic
◯ Chad
◯ Chile
◯ China
◯ Colombia
◯ Comoros
◯ Congo Democratic Republic of the
◯ Congo Republic of the
◯ Costa Rica
◯ Cote d'Ivoire
◯ Croatia
◯ Cuba
◯ Cyprus
◯ Czech Republic
◯ Denmark
◯ Djibouti
◯ Dominica
◯ Dominican Republic
◯ East Timor
◯ Ecuador
◯ Egypt
◯ El Salvador
◯ Equatorial Guinea
◯ Eritrea
◯ Estonia
◯ Eswatini
◯ Ethiopia
◯ Fiji
◯ Finland
◯ France
◯ Gabon
◯ Gambia
◯ Gaza Strip
◯ Georgia
◯ Germany
◯ Ghana
◯ Greece

◯ Grenada
◯ Guatemala
◯ Guernsey
◯ Guinea
◯ Guinea-Bissau
◯ Guyana
◯ Haiti
◯ Honduras
◯ Hong Kong SAR
◯ Hungary
◯ Iceland
◯ India
◯ Indonesia
◯ Iran
◯ Iraq
◯ Ireland
◯ Israel
◯ Italy
◯ Jamaica
◯ Japan
◯ Jordan
◯ Kazakhstan
◯ Kenya
◯ Kiribati
◯ South Korea
◯ Kosovo
◯ Kuwait
◯ Kyrgyzstan
◯ Laos
◯ Latvia
◯ Lebanon
◯ Lesotho
◯ Liberia
◯ Libya
◯ Liechtenstein
◯ Lithuania
◯ Luxembourg
◯ Macau SAR
◯ Madagascar
◯ Malawi
◯ Malaysia
◯ Maldives
◯ Mali
◯ Malta
◯ Marshall Islands
◯ Mauritania
◯ Mauritius
◯ Mayotte
◯ Mexico
◯ Federated States of Micronesia
◯ Moldova
◯ Monaco
◯ Mongolia
◯ Montenegro
◯ Morocco
◯ Mozambique
◯ Myanmar
◯ Namibia
◯ Nauru
◯ Nepal
◯ The Netherlands
◯ New Zealand
◯ Nicaragua
◯ Niger
◯ Nigeria
◯ North Macedonia
◯ Norway
◯ Oman
◯ Pakistan
◯ Palau
◯ Panama

◯ Papua New Guinea
◯ Paraguay
◯ Peru
◯ Philippines
◯ Poland
◯ Portugal
◯ Qatar
◯ Romania
◯ Russia
◯ Rwanda
◯ Saint Kitts and Nevis
◯ Saint Lucia
◯ Saint Vincent and the Grenadines
◯ Samoa
◯ San Marino
◯ Sao Tome and Principe
◯ Saudi Arabia
◯ Senegal
◯ Serbia
◯ Seychelles
◯ Sierra Leone
◯ Singapore
◯ Slovakia
◯ Slovenia
◯ Solomon Islands
◯ Somalia
◯ South Africa
◯ South Sudan
◯ Spain
◯ Sri Lanka
◯ Sudan
◯ Suriname
◯ Sweden
◯ Switzerland
◯ Syria
◯ Taiwan
◯ Tajikistan
◯ Tanzania
◯ Thailand
◯ Timor-Leste
◯ Togo
◯ Tonga
◯ Trinidad and Tobago
◯ Tunisia
◯ Turkey
◯ Turkmenistan
◯ Tuvalu
◯ Uganda
◯ Ukraine
◯ United Arab Emirates
◯ United Kingdom
◯ United States
◯ Uruguay
◯ Uzbekistan
◯ Vanuatu
◯ Vatican City
◯ Venezuela
◯ Vietnam
◯ Yemen
◯ Zambia
◯ Zimbabwe

---

How many years are you involved in PCD care/research?

◯ Less than 5 years
◯ 5-10 years
◯ Over 10 years

| | |
|---|---|
| Are you | ◯ Female<br>◯ Male<br>◯ Other/prefer not to disclose |
| Have you led or participated in PCD research project(s) during the past 15 years? | ◯ Yes<br>◯ No |
| What is your main area of interest related to clinical/epidemiological PCD research? | ______________________________<br>(please shortly describe) |
| Have you ever applied for funding for a PCD-related research project? | ◯ Yes<br>◯ No |
| How is PCD research predominantly funded at your centre/institute? (Tick all that apply) | ☐ Governmental/institutional funding<br>☐ Competitive grant funding (national or European, international)<br>☐ Funds from smaller foundations and charities, including patient support groups<br>☐ Funds from another institute through collaborative research<br>☐ Most research in my institute is not funded |
| Do you consider obtaining funding for PCD research is (select what applies best) | ◯ More difficult than other respiratory diseases or other diseases in my field<br>◯ Easier than other respiratory diseases or other diseases in my field<br>◯ I see no difference compared to other respiratory diseases or other diseases in my field<br>◯ I don't know |

**Please rate on a scale of 1 (not relevant) to 5 (very relevant) your opinion on whether the following factors are important or not important barriers to obtaining research funding for clinical and epidemiological PCD research:**

| | |
|---|---|
| PCD is too rare | *(Place a mark on the scale above)* |
| PCD is not considered severe/low mortality rate | *(Place a mark on the scale above)* |
| Lack of awareness about PCD in the medical community and public | *(Place a mark on the scale above)* |
| Lack of expertise/low impact publication record of the applicant research team | *(Place a mark on the scale above)* |
| Lack of potential commercial applications of the research | *(Place a mark on the scale above)* |

Research funding is very competitive

(Place a mark on the scale above)

Funders are more interested in basic science research projects

(Place a mark on the scale above)

Lack of evidence to support hypothesis/ lack of existing framework or research tools (e.g. datasets)

(Place a mark on the scale above)

Lack of local support in drafting a funding application (e.g. clinical research unit)

(Place a mark on the scale above)

**In addition to funding, which of the following factors are an additional challenge in performing PCD research at your institute/country? Please rate them on a scale of 1 (not relevant) to 5 (very relevant):**

Small number of patients

(Place a mark on the scale above)

Absent, small or inactive patient support group

(Place a mark on the scale above)

Heterogeneity of PCD diagnosis and clinical presentation

(Place a mark on the scale above)

Lack of dedicated research time for clinicians/ large clinical load

(Place a mark on the scale above)

Lack of good local or extended collaborative network

(Place a mark on the scale above)

Lack of interest from most colleagues

(Place a mark on the scale above)

Small number of team members with expertise in the field of PCD

(Place a mark on the scale above)

Patients are not motivated to participate (i.e. many research projects, patients live too far away etc.)

(Place a mark on the scale above)

Lack of resources (e.g. equipment, databases)

(Place a mark on the scale above)

**Please rate on a scale of 1 (strongly disagree) to 5 (strongly agree) the following statements considering PCD-related clinical and epidemiological research**

Most research projects require national or international multidisciplinary collaboration to achieve meaningful results.

(Place a mark on the scale above)

National and multinational registries and cohort studies are critical tools for epidemiological and clinical research..

(Place a mark on the scale above)

Patient support groups can have an important role in funding research, encouraging patients to participate and disseminating research results.

(Place a mark on the scale above)

Involvement of patients in all different stages of a research project can lead to better and more meaningful research.

(Place a mark on the scale above)

Standardisation of care and of clinical information collected may contribute to improvement of data quality and therefore to better research.

(Place a mark on the scale above)

**Please rank the following questions on research priorities related to PCD diagnosis, in order from 1 (most important) to 3 (least important):**

| | 1 | 2 | 3 |
|---|---|---|---|
| Who should undergo PCD diagnostic testing? | ○ | ○ | ○ |
| How to improve the accuracy, speed, and cost-effectiveness of diagnostic testing in different age groups and health care settings? | ○ | ○ | ○ |

| | | | |
|---|---|---|---|
| What is the true prevalence of PCD in different regions and the proportion of patients misdiagnosed as other respiratory diseases? | ○ | ○ | ○ |

**Please rank the following questions on research priorities related to PCD presentation, prognosis, and follow-up, in order from 1 (most important) to 4 (least important):**

| | 1 | 2 | 3 | 4 |
|---|---|---|---|---|
| How are upper and lower airways disease associated in patients with PCD? | ○ | ○ | ○ | ○ |
| What is the clinical variability and natural course of upper and lower respiratory disease in PCD, and which factors affect disease prognosis? | ○ | ○ | ○ | ○ |
| How should we best monitor disease progression in different age groups and health care settings? | ○ | ○ | ○ | ○ |
| Which health-related behaviours or everyday interventions can have a positive role in improving symptoms or quality of life in people with PCD? | ○ | ○ | ○ | ○ |

**Please rank the following questions on research priorities related to PCD treatments, in order from 1 (most important) to 5 (least important):**

| | 1 | 2 | 3 | 4 | 5 |
|---|---|---|---|---|---|
| Which of the already available and currently used medication and other management approaches for upper and lower airways are suitable for PCD patients? | ○ | ○ | ○ | ○ | ○ |
| How should we manage PCD in different age groups including asymptomatic or pre-symptomatic patients? | ○ | ○ | ○ | ○ | ○ |
| Which are the most suitable clinical and patient-reported outcomes to be used as endpoints in PCD clinical trials focused on the upper and lower airways? | ○ | ○ | ○ | ○ | ○ |

| | | | | | |
|---|---|---|---|---|---|
| Are there any genetic or molecular treatments in the pipeline that could help restoring ciliary function? | ○ | ○ | ○ | ○ | ○ |
| How could we improve care for specific patient categories such as adolescents, pregnant women, adults with comorbidities, and minority groups? | ○ | ○ | ○ | ○ | ○ |

**Please rank the following questions on other research priorities related to PCD, in order from 1 (most important) to 3 (least important):**

| | 1 | 2 | 3 |
|---|---|---|---|
| How is fertility affected in patients with PCD and what are the best fertility management approaches? | ○ | ○ | ○ |
| How is the mental health affected in people with PCD and their families? | ○ | ○ | ○ |
| How can we increase awareness and engagement of clinicians and patients in PCD research? | ○ | ○ | ○ |

**Please rank 3 of the following questions as the top 3 overall research priorities for PCD-related clinical and epidemiological research based on your opinion:**

| | your first priority | your second priority | your third priority |
|---|---|---|---|
| Who should undergo PCD diagnostic testing? | ○ | ○ | ○ |
| How to improve the accuracy, speed, and cost-effectiveness of diagnostic testing in different age groups and health care settings? | ○ | ○ | ○ |
| What is the true prevalence of PCD in different regions and the proportion of patients misdiagnosed as other respiratory diseases? | ○ | ○ | ○ |
| How are upper and lower airways disease associated in patients with PCD? | ○ | ○ | ○ |

What is the clinical variability and natural course of upper and lower respiratory disease in PCD, and which factors affect disease prognosis?

How should we best monitor disease progression in different age groups and health care settings?

Which health-related behaviours or everyday interventions can have a positive role in improving symptoms or quality of life in people with PCD?

Which of the already available and currently used medication and other management approaches for upper and lower airways are suitable for PCD patients?

How should we manage PCD in different age groups including asymptomatic or pre-symptomatic patients?

Which are the most suitable clinical and patient-reported outcomes to be used as endpoints in PCD clinical trials focused on the upper and lower airways?

Are there any genetic or molecular treatments in the pipeline that could help restoring ciliary function?

How could we improve care for specific patient categories such as adolescents, pregnant women, adults with comorbidities, and minority groups?

How is fertility affected in patients with PCD and what are the best fertility management approaches?

| | | | |
|---|---|---|---|
| How is the mental health affected in people with PCD and their families? | ◯ | ◯ | ◯ |
| How can we increase awareness and engagement of clinicians and patients in PCD research? | ◯ | ◯ | ◯ |


projectredcap.org
